# Do Men and Women “Lockdown” Differently? An Examination of Panama’s COVID-19 Sex-Segregated Social Distancing Policy

**DOI:** 10.1101/2020.06.30.20143388

**Authors:** Liana R. Woskie, Clare Wenham

**Author notes:** co-chair of the Gender and COVID Working Group. **CORRESPONDING AUTHOR:** Clare Wenham, Ph.D., Assistant Professor of Global Health Policy, Department of Health Policy, London School of Economics, Houghton Street, London, WC2A 2AE.

## Abstract

Mobility enables individuals to generate income and is a key input for empowerment and personal autonomy. Curtailment of aggregate social mobility - through policies such as: social distancing recommendations, shelter in place orders and state-enforced lockdowns - has become a primary strategy to address COVID-19 to limit social contact and reduce disease transmission. In this context, a small number of countries have instituted gender or sex-segregated mobility policies (Panama, Peru, and Bogota, Colombia). Through a retrospective analysis of global geographic positioning (GPS) data, this study presents an overview of aggregate mobility in Panama following the country’s implementation of a sex-segregated social distancing policy. Panama was selected as the nationwide sex-segregated policy was enforced throughout the lockdown period. The paper looks at mobility trends on female- and male-sex mobility days, examining differences by volume of movement and type of community locations visited as compared to pre-COVID trends. We find lower visits to all community location categories on female-mobility days. However, we found no significant difference in visits to “workplace” locations on male- v. female-mobility days. The paper discusses the implications of these findings in three areas: (1) Informal burden of labor and social reproduction, (2) Implications for women’s autonomy and safety in the home, and (3) Women’s economic empowerment. In addition, it raises open ethical questions regarding gender identity in COVID-19 policies.

## INTRODUCTION

Panama adopted one of the most aggressive responses to COVID-19 in Latin America; scoring highly on Oxford’s COVID-19 policy response stringency index (Anon 2020). This has centered on a lockdown that allows for mobility based on an individual’s sex, as listed on their national identification card *“cedula”* (Anon 2020).^1^ Accordingly, on Mondays, Wednesdays and Fridays, women can leave their homes while on Tuesdays, Thursdays and Saturdays only men can. The policy was justified to reduce the volume of individuals on the street at any one time and, in doing so, reduce the risk of disease transmission.

Beyond describing social distancing policies that have emerged in response to COVID-19, there is an increasing urgency to better understand their impact, particularly their differential effects across a population. Early (albeit incomplete) sex-disaggregated data has shown differences in vulnerability to COVID-19 (Anon 2020): more men than women are dying, potentially due to sex-based immunological difference (Chen et al. 2020) or – increasingly likely – social factors, such as patterns and prevalence of smoking, and other demographic factors unrelated to biological sex (Liu et al. 2017). In line with this, early research from the United States suggests that men may be less likely to undertake certain public health precautions, such as engaging in social distancing practices or hand-washing (Anon 2020). However, corresponding data on outcomes are incomplete and reliant on national testing strategies which vary in their accessibility, making early assessments of sex-disaggregated death from COVID-19 imprecise (Wenham, Smith, and Morgan 2020).

Apart from mortality, there may be primary and secondary gendered impacts of epidemic disease and associated response. Previous research has demonstrated women can have a higher risk of exposure to infectious disease due to formal care giving (70 percent of the global healthcare workforce are women) and informal care within families, which has been mirrored in early testing statistics for COVID-19 (Harman 2016, Anon 2020). Women have disproportionately experienced downstream effects of prior outbreaks and policies enacted to mitigate risk (Wenham, forthcoming). This includes increased domestic work for disease control interventions (quarantine, vector control, (Wenham, Nunes, et al. 2020) additional labor of childcare and home schooling (Anon 2020), interruption of routine maternal health provision during epidemics (Anon 2020) and increased economic insecurity owing to higher precarious employment contracts (Bandiera et al. 2019). Despite the existing literature interlinking gender and outbreaks, we lack a comprehensive understanding of the gendered impacts of the widespread curtailment of human mobility undertaken in response to COVID-19.

The sex-segregated policy enacted in Panama and newly available data on mobility across the country provide a unique opportunity to examine early differences in aggregate mobility during COVID-19 on male-versus female-mobility days and what this may reveal about norms of social reproduction, paid and unpaid labor and how people interact during lockdown. We therefore answer the following questions: First, is there a significant difference in aggregate mobility to community locations between male-versus female-mobility days? Second, how does the change in aggregate mobility compare by location category? Finally, what are potential implications of these differences on social reproduction, division of labor or women’s autonomy?

## POLICY CONTEXT

On March 30, 2020 the Panamanian Ministry of Public Security launched “Executive Decree 507” detailing the country’s sex-segregated mobility policy to reduce civic circulation, stating: “*From Wednesday 1*^*st*^ *April, public mobility will be conditional on sex as stated on national identification documents {cedula}*,”(Anon 2020). The logic motivating this policy was that it provided a relatively simple way to reduce public circulation by half, and thus easily enforce social distancing in public spaces (Anon, 2020). Mobility was further limited by the final number listed on each *cedula* which dictated the hours you could enter supermarkets and pharmacies. Every Panamanian citizen, naturalized citizen and permanent resident has a *cedula* identification card, and is obliged to carry it in public. Thus it was considered the most “equitable” way to easily control public mobility rapidly. Juan Pinto, Minister for Public Security stated “*This [policy] is for nothing more than to save your life*,” (Anon, 2020).

Notably, the policy was based on an individual’s sex, as listed on their *cedula*. The binary distinction as the basis of a national lockdown, a movement-restricting policy, raised immediate concerns amongst transgender and otherwise non-binary communities (Anon, 2020). However, despite widespread domestic and international concern, the policy remained sex-segregated based on the *cedula* throughout its duration.

Following its announcement on March 30, Panama’s policy was replicated by the government of Peru and the city of Bogota, recognizing the easy enforceability of such a policy and the impact it could have on reducing public circulation. However, on April 10, 2020 (within 8 days of its enactment) Peru’s government cancelled their ‘*pico y genero*” (peak [hours] and gender) policy; with President Marin Vizcarra announcing that the policy had been insufficient, and failed in its objectives. This was later qualified with additional commentary on the decision to enact a sex-segregated policy more generally, “In our patriarchal world, there are roles assigned to women which must be challenged, but now [amid pandemic] is not the time to fight them,” (Anon 2020). Meanwhile in Bogota, the mayor launched a similar gender-based policy on April 13, 2020 dividing women and men’s days not by weekdays, but by odd and even dates in the month.

## THEORETICAL FRAMEWORK

### Sex, Gender and Infectious Disease

The intersection of sex and infectious disease has been documented. Early social determinants of health work show that sex influences health (Marmot 2005). While sex and gender are often conflated in this literature, research shows that women face a double burden of infectious disease (Lee and Frayn 2008), being those most exposed through formal and informal care work with patients. This risk of infection appears be driven by social factors compound by gender norms i.e. that women further undertake the majority of “work” involved in caregiving and response efforts, as opposed to biological sex. This can range from being responsible for contraception to limiting sexual transmission,(Brown 2015) to vector control efforts (Wenham, Nunes, et al. 2020), or managing the additional childcare and domestic responsibilities when schools shut, and quarantines are implemented (Wenham, Smith, et al. 2020).

Prior to COVID-19 the intersection between gender and infectious disease had rarely been considered by policymakers, albeit documented in the academic literature (Wenham, forthcoming; Davies and Bennett 2016; Harman 2016). In addition, neither sex nor gender have been meaningfully mainstreamed into policy to respond to infectious disease (Smith 2019). Thus, this sex-segregated mobility policy in Panama is unusual in considering sex a mechanism of disease control. However, the division of sexes for this policy was not based on the inherent differential effects of the outbreak or to redress the burden of response efforts. Instead sex-stratification was a simple mechanism utilized by the Panamanian government to rapidly reduce the number of people circulating by theoretically halving the quantity each day. Regardless, due to implementation based on sex, the Panama policy raises questions regarding the differential effect of mobility curtailment policies between the two groups and potential effects such a policy may have on broader issues of equality.

### Social Reproduction

In line with this, we consider the implications of the Panama policy on social reproduction. Across political spectrums, gendered norms and activities are often dictated by everyday political economy (Sjoberg 2016). Social reproduction includes those household activities central to production and reproduction of life and capital economic contribution (Anon 2020). Social reproduction includes, but is not limited to: childrearing, caring responsibilities, small-scale agricultural labor, household work and maintenance. This is not to suggest that all women have identical roles across houses, communities and the globe, but social reproduction recognizes global patterns of informal, often invisible or devalorized work, which is usually carried out by womxn, regardless of the role it plays in capital development.

Feminist economists have demonstrated how this invisible ‘feminized’ labor within the private home is a fundamental lynchpin facilitating others, notably men, in their contribution to the public, paid, workforce, and therefore capitalist global economy. In this way, value of this feminized labor is vital to the capitalist functioning system. Smith (1990) argues that the conceptual division between the female private space and the public male space maintains the dominance of men in the practice of globalizing, gendered capitalism, and thus societal and global power (Smith 1990). Conversely a global capitalist system can have downstream effects on this feminized social space; economic crises create significant impact on (social) reproduction, as demonstrated by disease outbreaks (Elson 1994; Roberts 2013). During the Ebola outbreak women who lost their jobs were out of work for longer than men the wake of the crisis. Similarly, girls were out of school for longer (Bandiera et al. 2019). For Zika, social reproduction was demonstrated in women performing vector control activities, but furthermore if they had a child borne with Congenital Zika Syndrome they had to leave their jobs to provide full time care for those children, and most were abandoned by their partners and thus this care activity was left solely to women (Wenham, Forthcoming) (Diniz and Grosklaus Whitty 2017). Understanding the intersection between social reproduction and mobility can shed a light on women’s agency within political and economic systems, and how outbreaks can affect women’s economic empowerment.

### Physical Autonomy and Mobility

In recent models, mobility (or physical autonomy) has consistently been identified as a primary dimension of women’s autonomy (Jejeebhoy and Sathar 2004, Osamor and Grady 2016, Samari and Pebley 2018). Control over one’s life, or “autonomy” is viewed as a set of inter-linked domains. Physical autonomy can be defined as an individual’s ability to freely interact with the outside world, or the extent to which an individual is free of constraints on their physical mobility (Jejeebhoy and Sathar 2004). In addition to its intrinsic value, physical autonomy is also instrumentally essential. Mobility is fundamental to livelihoods, everyday life, communities, and individuals. Accordingly, research has demonstrated how mobility, or lack thereof, has gendered implications and indeed molds gendered assumptions. For example, tracing women’s movements in the public sphere can demonstrate a move towards women’s economic opportunity and social empowerment, and often higher wages and challenging traditional gender power structures (Hapke and Ayyankeril 2004; Mandel 2004). Conversely, limitations of mobility reproduce notions of a public/private divide, with gender norms often placing women in the home (Hanson 2010; Sager 2016).

However, mobility is challenging to assess. It is common for indices that measure physical autonomy to sum the number of places to which a woman can go unescorted. For example, Jejeebhoy’s 2001 index used to assess autonomy in India and Pakistan ranged from 0 (if a woman must be escorted everywhere) to five (if she could move unescorted to five select location categories)(Jejeebhoy and Sathar 2004). Data to populate these indices has been collected from women themselves in the form of household survey tools, posing questions regarding the ability to engage in a range of activities. Overall mobility, however, is rarely included; it is challenging to measure from self-report in a consistent or meaningful way across populations. Thus, aggregate changes in mobility are rarely included in estimates of autonomy. The unique scenario of a COVID-19 sex-segregated mobility policy, paired with recently accessible GPS data, allows us to look at trends in mobility and, in turn, disparities in mobility by sex; a critical yet previously under-explored aspect of autonomy.

## METHODS

In this paper we take a novel approach to assessing curtailed mobility by looking at changes in aggregate social mobility, or visits to community locations, over time. To do so, we conducted a retrospective analysis of Global Positioning System (GPS) data from Panama between February 15 and May 29, 2020 (the last weekday that the policy was in effect). We obtained aggregated anonymized data from Google’s COVID-19 Community Mobility Reports, a dataset made up of Google users with mobile devices across Panama. Users opted in to sharing their Location History, a feature which is turned off by default (Aktay et al. 2020). The anonymized dataset resulting from these devices is aggregated daily. It is the same data used to create publicly-available Google COVID-19 Community Mobility Reports, the aggregation and anonymization process has been previously described (Aktay et al. 2020).

We look at the relative change in the number of visits to community locations; specifically, relative change in the average number of visits to Google’s five non-residential location categories: 1) grocery and pharmacies, 2) retail stores, recreational sites, and eateries, 3) transit stops, 4) parks and 5) workplaces. For each location category studied, each location history user can contribute at most once to each category (Aktay et al. 2020). To look at these five variables, we use Google’s standard algorithm to assess how visits and length of stay at different locations change compared to a baseline. In this model, changes for each day are compared to a baseline value for that day of the week. The baseline is the median value, for the corresponding day of the week, during the 5-week period from January 3rd to February 6th 2020 (Aktay et al. 2020). For example, if a value is −70 on a Monday, it is 70 percent lower than a “baseline” value based on prior mobility on Mondays during the five-week pre-period. Importantly, for our analysis, during the Panamanian lockdown, many locations falling within categories two and four (as defined by Google, Appendix Table 1) were formally closed, with location three (transit stops) largely available for those with a *salvoconducto* or going to grocery and pharmacy stores. The only workplaces which were open (location five) were those providing essential services.

Conceptually, we are comparing male- and female-mobility days to mobility trends prior to COVID-19 when people of all genders could move freely. Because the curtailment of overall mobility is an effort to slow the spread of the COVID-19 pandemic and may be positive, our primary interest is the differential effect of curtailment on male-v. female-mobility days. For this reason, we perform simple two-sided t-tests on the equality of means (Table 2) to assess if there was a meaningful difference in the change in mobility between the two types of days as compared to the non-gender segregated baseline.

**Table 1.** Timeline of Panama’s Policies and National Movement Restrictions in Response to COVID-19.

|  |  |
| --- | --- |
| March 12 | Schools and workplaces closed |
| March 22 | The Panamanian government suspends all international commercial flights for 30 days. |
| March 25 | Panama enacts nationwide movement restrictions, (quarantine). The movement restrictions will include regular windows of time for people to conduct essential activities, such as grocery shopping, getting gas and medicines, or dealing with a medical emergency, based on the last digit of their cedula number or passport if an individual does not have a *cedula*. |
| March 26 | Panama announces nationwide suspension of commercial passenger and domestic charter flights, in addition to the March 22 ban on international commercial passenger flights. There are exceptions for cargo, humanitarian, medical supplies, medical evacuation, and vaccines. |
| March 30 | The government releases "Executive Decree 507" announcing expanded movement restrictions based on gender. Individuals are permitted to circulate during the same hours determined by *cedula* as set forth in previous decrees. Sales of alcohol were prohibited to limit criminal behavior and/or violence |
| April 1st | The government enacts gender restrictions, with the following daily designations:<br>- Women: Monday, Wednesday, and Friday<br>- Men: Tuesday, Thursdays, and Saturday<br>- Exceptions apply for holders of permission letters (*salvoconductos*) |
| April 21 | The Municipal Council of the City of Panama passes a new decree stating that anyone leaving their residence must be wearing a mask that covers their nose and mouth. Panamanian National Police and Municipal Police will enforce the decree which extends throughout the metropolitan area of Panama City. |
| April 23 | The Government of Panama announces that both Saturday, April 25, and Sunday, April 26, may be full quarantine whereby no one can leave their homes. |
| May 13 | Select re-opening / mobility permissions extended for workforce, such as: mechanics, plumbers, electricians, air condition repairs, pool repairs, fisherman etc. |
| May 27 | End of the lockdown announced, including relaxation of limitations based on *cedula*. |
| June 1 | National quarantine relaxed, based on a 6 stage process detailed in the policy "Return to Normality" |

**Table 2.** Differences in Aggregate Mobility on Sex-segregated Mobility Days by Location Category.

|  | Male-Mobility Weekdays |  | Female-Mobility Weekdays |  | p-value |
| --- | --- | --- | --- | --- | --- |
|  | Mean | 95% CI | Mean | 95% CI |  |
| Change in Aggregate Mobility Through May 13<sup>th</sup> |  |  |  |  |  |
| Retail and Recreation | -73.4 | -75.7 – -71.1 | -77.8 | -80.2 – -75.4 | 0.011 |
| Grocery and Pharmacy | -41.5 | -43.9 – -39.0 | -54.6 | -58.0 – -51.2 | 0.000 |
| Transit Stations | -71.9 | -73.6 – -70.3 | -75.5 | -77.6 – -73.5 | 0.012 |
| Parks | -64.8 | -66.9 – -62.8 | -68.9 | -71 – -66.8 | 0.008 |
| Workplace | -70.2 | -72.6 – -67.8 | -71.4 | -74.5 – -68.3 | 0.549 |
| Change in Aggregate Mobility Through the End of the Policy |  |  |  |  |  |
| Retail and Recreation | -71.4 | -73.6 – -69.2 | -75.3 | -77.6 – -72.9 | 0.025 |
| Grocery and Pharmacy | -40.9 | -42.7 – -39.2 | -51.9 | -54.9 – -48.9 | 0.000 |
| Transit Stations | -69.9 | -71.9 – -67.8 | -72.8 | -75.1 – -70.5 | 0.073 |
| Parks | -62.5 | -64.9 – -60.1 | -66.3 | -68.6 – -63.9 | 0.032 |
| Workplace | -67.1 | -70.1 – -64.0 | -67.8 | -71.1 – -64.6 | 0.734 |

Notes: all p-values are two tailed; resulting from a two-sample t-test assessing the difference in means between male v. female-weekdays

## RESULTS

We find a substantial drop in mobility from the start of the outbreak, which was sustained after the lockdown was implemented on April 1, 2020 with notable weekly variation in mobility. Analysis of different days and location categories, those with the least change from the pre-COVID baseline are grocery and pharmacies. This category remains that with the least change from baseline over the lockdown period, potentially keeping with the strict enforcement of all retail and other service industries being closed.

**Figure 1.**
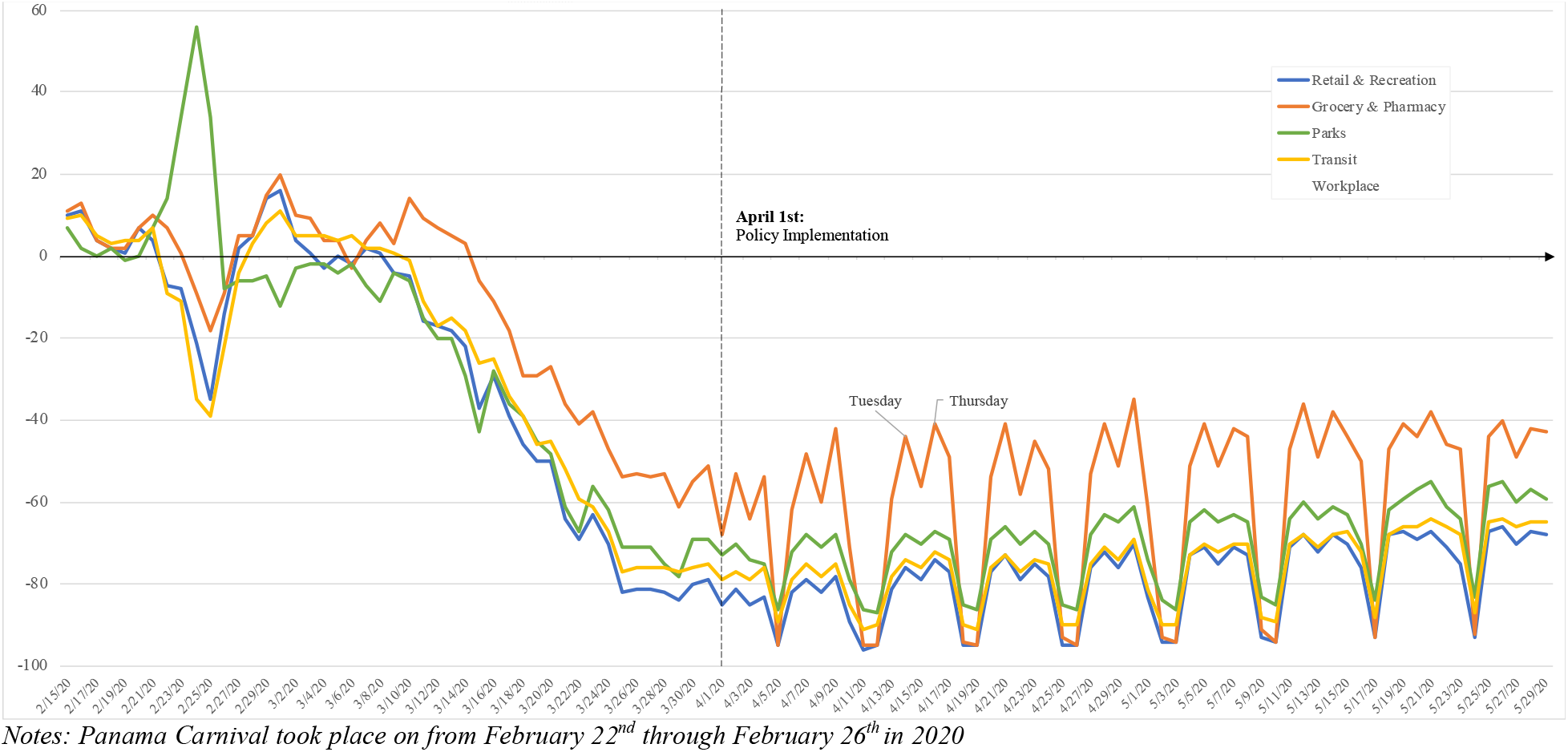
Trends in Aggregate Mobility to Community Locations Across Panama Over Time.

However, our data demonstrate that there was less of a change in aggregate mobility from the baseline for grocery and pharmacies on “male” days compared to female days (Table 2). In fact, for all mobility categories, aggregate male-day mobility was closer to the baseline than female-day mobility, with a 95% confidence interval. In addition, while visits to all categories remained low when compared to the baseline, they increased over time with slopes ranging from 0.29 (transit) to 0.38 (grocery and pharmacy), Appendix Figure 1. Extending our timeframe to the end of the policy, we find that the difference in means for aggregate mobility to transit stations is no longer statistically significant between male- and female-mobility days.

In addition, there was no statistically significant difference in “workplace” aggregate mobility between male and female days. Given that most of the labor market was suspended, or asked to work at home, workplace mobility in theory refers to those considered essential workers involved in the COVID-19 response and/or vital infrastructure services across genders, whose mobility was not sex-segregated. Individuals traveling to workplaces were provided with exemption letters *salvoconductos* to facilitate transit.

## DISCUSSION

We find that Panama’s sex-segregated lockdown resulted in significantly different aggregate mobility on male-v. female-mobility days as compared to a pre-COVID baseline, suggesting that men and women may be experiencing the “lockdown” differently. Differences were observed in both the volume and type of locations visited; with lower visits to all community location categories on female-mobility days. This difference was particularly pronounced in visits to grocery and pharmacy locations. This was at odds with media accounts from Peru highlighting longer queues outside supermarkets on “female” days, and assumptions of the gendered distribution of domestic labor (Anon 2020). These findings motivate further discussion of the impact of sex-segregated policy on women’s curtailed mobility. It appears women may not be performing certain types of informal labor; specifically, they are undertaking fewer tasks outside the home. Indeed, given men’s mobility was closer to the pre-COVID baseline they appear to be visiting grocery shops and pharmacies more frequently on their permitted days. This finding suggests that men may be performing public domestic activities within families and/or communities at higher rates than women. Further investigation is required to understand these trends and the extent to which this differs from normal mobility patterns, if and why men are undertaking these activities and, conversely, why women are not. Below, we discuss potential implications for social reproduction, women’s autonomy and safety in the home and economic empowerment.

### 1. Social Reproduction

During COVID-19, social reproduction is evident. Women are performing the majority of the care in hospitals and care homes (Anon 2020). Moreover, and in line with our findings, time use surveys and polling has demonstrated that women are performing the majority of the domestic burden; caring for the children who are not in school, undertaking additional housework associated with the increased time in the home, and are more anxious than men about the lockdown and associated disruptions (Anon 2020). Compounding this, early data suggest that during COVID-19 women are more likely to have been furloughed or made redundant as they are more likely to be on part-time, flexible or zero hours contracts, or employed in the informal sector (Anon 2020, Phimister et al. 2020).

Our work suggests that women may be experiencing differential rates of curtailed mobility during lockdowns in ways that appear to reaffirm social reproduction in some ways and not in others, posing questions about mobility, autonomy and women’s agency to undertake tasks outside the home. Given overall lowered mobility across society, tasks outside the home during lockdown may be ascribed new value during lockdown which in turn impacts household bargaining (Agarwal 1997). For example, going to the grocery store may be one of the only opportunities to leave the home and engage with non-household members. It may also be that men are “sent” to the shops or choose to seek respite from the private domain. While it may have previously been valued as a purely domestic task, it now provides novel social benefit reflected in household negotiations. This reaffirms the invisibility of women even within their own domestic spaces. Alternatively, the in-home domestic burden borne by women may be high enough that they are unable to or may not have the agency to engage in external tasks. Men are “going to the store” but this could be less a pro-active bargaining choice on their part than a reflection of women’s burden in the home, e.g. undertaking childcare. A final consideration might be gendered perception of risk, and whether going out in public and potentially being exposed to infection is evaluated differently by men and women. Our data do not reveal the motivating factor behind men’s differential mobility comparable to women, but they raise questions about women’s autonomy and mobility, reproduce notions of the public/private divide, and suggest that women may self-isolate to a greater degree than men (whether an active choice or not). This is an important finding for infectious disease control interventions. Yet we do not suggest that such a policy should be replicated, as we are yet to understand the downstream effects of sex-segregated isolation, and how such policies may disenfranchise women and jeopardize their physical and economic security.

### 2. Implications for Women’s Autonomy and Safety in the Home

These findings raise concern regarding women’s mobility in Panama. In the short term, the lack of mobility might optimistically mean that women are abiding by the strict quarantine measures, that they are simply “better at social distancing” and thus are less likely to be exposed to COVID-19. This may be the case, but unequal rates of curtailed mobility raise questions regarding women’s physical autonomy in the near term. This has immediate practical implications e.g. concerns over foregone access to food, access to healthcare, etc. Food shortages, whether caused by supply or demand side, are increasingly apparent in COVID-19 (Anon 2020, Anon 2020). Similarly, limitations of health service providers to only offer “essential services” beyond COVID-19 related care, as well as demand side concerns over infection risk of individuals in healthcare settings are increasingly evident in the outbreak. If women have the added burden of limited mobility, this may also impact health seeking behavior along gendered lines. Previous research demonstrated that, excluding obstetric care, women are less likely to visit hospitals, noting out-of-pocket expenditure, travel expenses and discrimination for travelling alone as reasons for low attendance (Anon 2020). Moreover, globally there have been trends of reduced admissions to emergency rooms during the pandemic, with medics suggesting this reflects individual’s concern about disease transmission in hospitals (Thornton 2020). We are yet to secure sex-disaggregated data for health seeking behavior during COVID-19 for non-pandemic related health concerns to understand how different perceptions and/or domestic demands may alter this interaction with the health system.

Lack of mobility also poses concerns with increased time in the home and narrowed social networks, which have historically compounded issues of intimate partner violence (IPV) and safety in the home (Lanier and Maume 2009)(Goldenberg et al. 2014; Pronyk et al. 2006). COVID-19 has amplified existing rates of IPV globally, with estimates of increases to calls to domestic violence hotlines increasing 60 percent in Europe (Mahase 2020) and alarmingly up to 79 percent in Colombia during March and April 2020 (Anon 2020). Notably, as part of the COVID response, the Panamanian government also prohibited the sale of alcohol during the quarantine period in an effort to reduce violence. Official statistics in Panama suggest a significant decrease in rates of such violence (Anon 2020), but further specialists have suggested that it is exactly this lack of mobility to report these crimes explain the low numbers (Mumtaz and Salway 2005). As IPV is under-reported, or data recorded through proxy measures after the fact, we must consider these data as they appear and include them in further analysis of the impact of sex-segregated policies for self-isolation.

### 3. Economic Empowerment

In the longer term, curtailed female mobility raises broader challenges for women’s economic empowerment. In the wake of the Ebola outbreak in West-Africa, women were out of work for longer than men in the post-crisis period (Bandiera et al. 2019). If women are not as mobile as men during the pandemic, this may extend into the post-COVID period and during the re-opening up of the economy. We have seen in Panama’s exit strategy that the first tranche of sectors opened are those which are traditionally male dominated; mechanics, construction, building maintenance and fishing (Anon 2020). Industries which traditionally employ women (education, hospitality, tourism) are delayed in re-opening until later in the “return to normal” government plan, placing further economic insecurity on women. Moreover, if women are performing the majority of the childcare, this will likely continue in the post-crisis period until schools re-open (not until stage five of the “return to normal” strategy) preventing women from returning to their jobs (Anon 2020). This can have widespread impacts on stability and economic development, each of which has been shown to improve with greater female participation in the labor market, as well as secondary effects on women’s civic and political participation. At time of writing, lockdown was still in place, and a number of sectors remain still closed, thus this remains speculative and we wait to see how economic empowerment of women is affected, and whether the government of Panama takes proactive steps to mitigate against the indirect effects on women posed by additional childcare, and the effects of which sectors of the economy are opened and in what order.

### 4. Gender-Identity

Finally, implementing a policy to limit mobility based on sex-segregation, presupposes that all individuals identify as male or female and that this identity aligns with the sex listed on their *cedula* as well as their gender presentation. With this assumption, this policy failed transgender and non-gender or sex-binary Panamanians from its inception. Early reports from Human Rights Watch suggest that transgender individuals, specifically, have suffered discrimination when leaving their homes on days that were in accordance with their gender identity, yet others for being outside of the home on days that were in accordance with the sex listed on the *cedula*: “*Transgender people in Panama are being humiliated and accused of breaking the law under the quarantine policy simply for being themselves*,” (Anon 2020). We require a deeper exploration of this issue; ensuring policies do not utilize citizens’ identities (or erasure of those identities) in ways that compound existing societal inequalities, particularly those borne out through law enforcement.

## LIMITATIONS

First, the data used is reliant on both the ownership of a smart phone and the decision to opt in to sharing data with Google. Because we lack information on individuals’ characteristics, we do not know the extent to which the population represented in Google’s aggregated data is reflective of the true population of Panama. In line with this, we do not have an estimation of how smart phone ownership may vary by gender, by socio-economic status or rurality. As a result, we only compare changes to the group’s pre-trend period. Second, we have no way to disaggregate men and women’s differential mobility prior to the policy’s implementation. While we compare aggregate mobility to a pre-COVID baseline, we do not know if the baseline reflects higher rates of mobility amongst men. Third, our analysis is only limited to data on weekdays. Although Saturdays were technically men’s days, they were subject to some additional mobility limitations throughout the study period. As a result, there is some chance that differences may be driven by men having fewer days. Finally, this study only utilizes data on mobility, which does not include information on the motivations for mobility and/or the activities undertaken whilst in public. For example, while we see increased visits to groceries and pharmacies on male-mobility days, we have no data on what was purchased on these days and whether this reflects differences in shopping trends.

## CONCLUSION

This data raises questions concerning the impact of sex-segregated mobility policies. First, it offers an indication that women may be adhering to lockdown to a greater extent than men, which may signal reduced risk of disease transmission. As sex-disaggregated data becomes more available, it is critical to explore this beyond Panama. Secondly, the difference in mobility on male- and female-mobility days provides early evidence that socialdistancing policies may differentially impact men and women. Women’s reduced mobility may be a reflection of“better social distancing,” but also causes concern regarding the distribution of labor (i.e. in dual parent households)and whether women may be less mobile due to reduced inter-household bargaining, or additional tasks they have undertaken within the home amplifying social reproduction. Third, more pronounced changes in aggregate mobility from baseline on female-mobility days raises concern for access to basic goods, such as food and healthcare, and if lockdown differences by sex may increase interpersonal violence. Fourth, mobility differences may impact women’seconomic empowerment in the medium-to long-term. Finally, the policy failed to recognize diverse gender identities and may reproduce inequalities and injustice for non-binary individuals with unknown long-term effects. More data, particularly utilizing mixed-methods, must be collected and collated to substantiate and better understand concerns raised.

## Data Availability

All data is publicly accessible.

https://www.google.com/covid19/mobility/

## APPENDIX TABLES AND FIGURES

**Appendix Table 1.** Google Mobility Location Categories.

|  | Example locations include: |
| --- | --- |
| *Retail and recreation* | Restaurants, cafes, shopping centers, museums, libraries and movie theatres |
| *Grocery and pharmacy* | Grocery markets, food warehouses, farmers markets, specialty food shops, drug stores and pharmacies |
| *Parks* | National parks, public beaches, marinas, dog parks, plazas and public gardens |
| *Transit Stations* | Public transport hubs, such as subway, bus and train stations, taxi stands |

More information on Google Location Categories can be found at on the COVID-19 Community Mobility Reports webpage: https://www.google.com/covid19/mobility/

**Appendix Figure 1.**
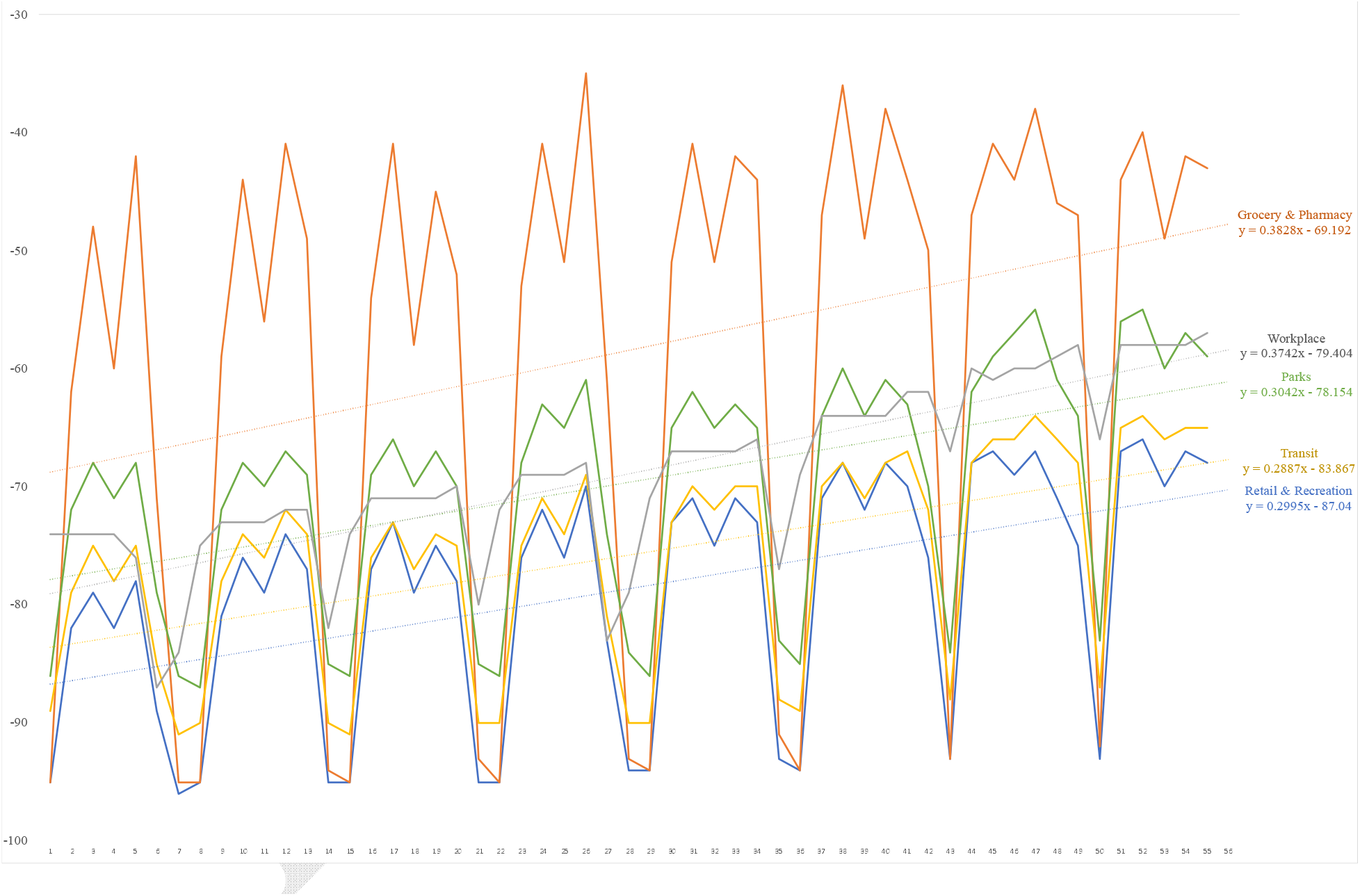
Trends in Aggregate Mobility Over Time by Location Category During the Sex-Segregated Quarantine.

**Appendix Figure 2 & 3.**
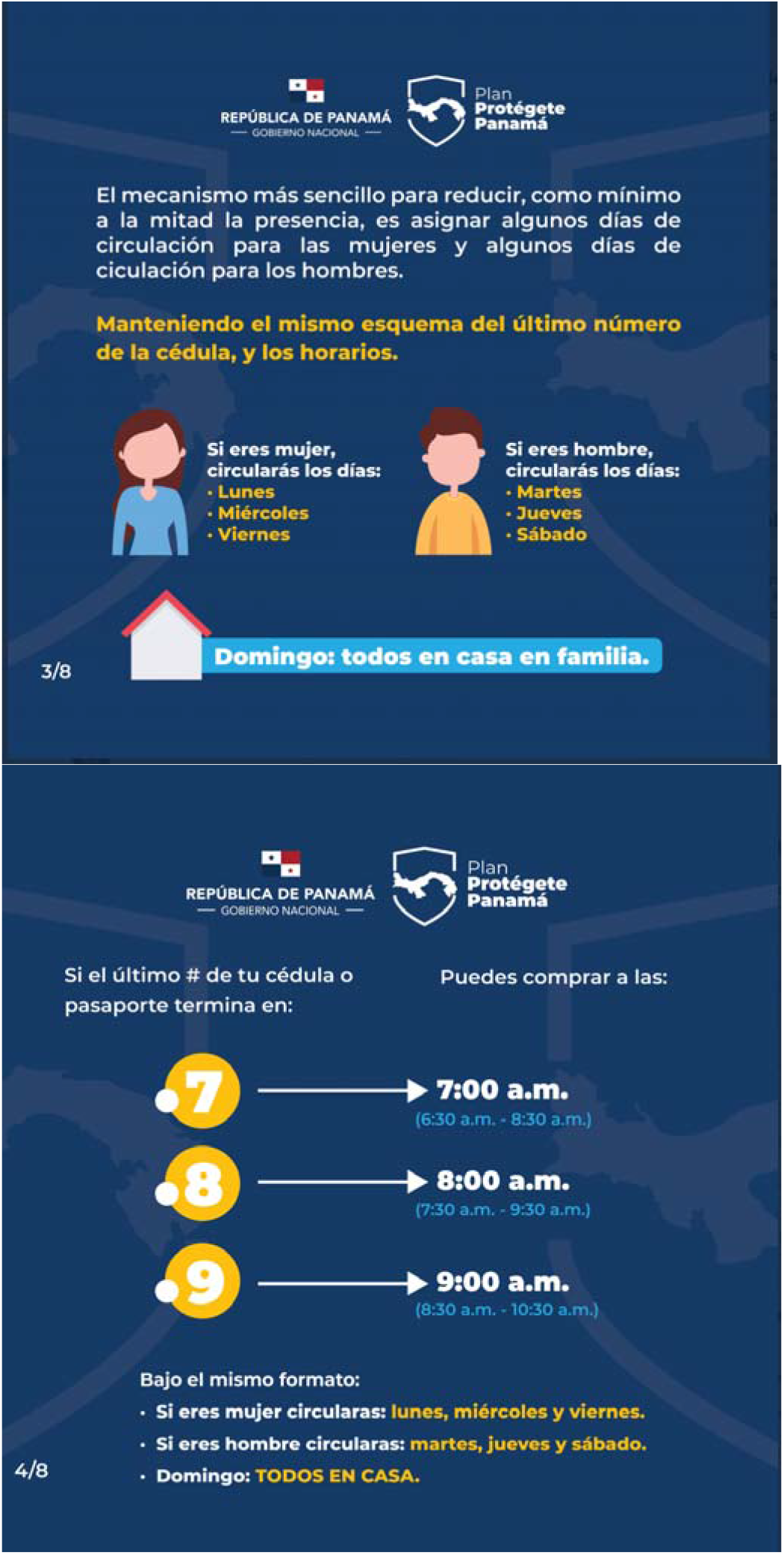
Description of Policy for Public, Republic of Panama.

## Footnotes

1 *The sex marker on the Panamanian ID card is based on an individual’s biological sex, a moniker assigned at birth based on the sex organs with which a Panamanian is born. The concept of sex is commonly understood as binary (male or female) and the policy was instituted as such. However, approximately two percent of the global population may be intersex or have a combination of sex organs. In addition, gender refers to the social attributes associated with being male or female. These attributes, and their attendant expectations, may or may not align with an individual’s sex, are socially constructed and can vary between contexts. Throughout this piece, we use the tAerms “men” and “women” as they are used within the national policy. Due to the spectrum of both sex and gender, this categorization is limited*.

## REFERENCES

Agarwal, Bina. 1997. “‘Bargaining’ and Gender Relations: Within and beyond the Household.” Feminist Economics 3(1):1–51.

Aktay, Ahmet, Shailesh Bavadekar, Gwen Cossoul, John Davis, Damien Desfontaines, Alex Fabrikant, Evgeniy Gabrilovich, Krishna Gadepalli, Bryant Gipson, Miguel Guevara, Chaitanya Kamath, Mansi Kansal, Ali Lange, Chinmoy Mandayam, Andrew Oplinger, Christopher Pluntke, Thomas Roessler, Arran Schlosberg, Tomer Shekel, Swapnil Vispute, Mia Vu, Gregory Wellenius, Brian Williams, and Royce J. Wilson. 2020. “Google COVID-19 Community Mobility Reports: Anonymization Process Description (Version 1.0).”

Anon. n.d. “[The Epidemiological Characteristics of an Outbreak of 2019 Novel Coronavirus Diseases (COVID-19) in China]. - PubMed - NCBI.” Retrieved April 14, 2020a (https://www.ncbi.nlm.nih.gov/pubmed/32064853).

Anon. n.d. “10 Key Issues in Ensuring a Gender Balance in the Global Health Workforce.” Retrieved April 20, 2020b (https://www.who.int/news-room/feature-stories/detail/10-key-issues-in-ensuring-gender-equity-in-the-global-health-workforce).

Anon. n.d. “Briefing: Covid-19 - Gender and Other Equality Issues - Womens Budget Group.” Retrieved June 4, 2020c (https://wbg.org.uk/blog/briefing-covid-19-and-gender-issues/).

Anon. n.d. Capítulo III-Delitos Contra La Identidad y Tráfico de Menores de Edad 3 2 1 0 0.

Anon. n.d. “Comunicado N° 35 MINSA COVID-19 – Ministerio de Seguridad de Panamá.” Retrieved June 1, 2020e (https://www.minseg.gob.pa/2020/03/comunicado-no-35-minsa-covid-19/).

Anon. n.d. “Counting Indirect Crisis-Related Deaths in the Context of a Low-Resilience Health System: The Case of Maternal and Neonatal Health during the Ebola Epidemic in Sierra Leone | Health Policy and Planning | Oxford Academic.” Retrieved April 2, 2020f (https://academic.oup.com/heapol/article/32/suppl_3/iii32/4621472).

Anon. n.d. “COVID-19: Jobs At Risk Index (JARI). Which Occupations Are Most at Risk? - Autonomy.” Retrieved June 5, 2020g (https://autonomy.work/portfolio/jari/).

Anon. n.d. “COVID-19 Survey Results | Altarum.” Retrieved June 1, 2020h (https://www.altarum.org/covid/results).

Anon. n.d. “Extensive Gender Discrimination in Healthcare Access for Women in India.” The BMJ. Retrieved June 4, 2020i (https://www.bmj.com/company/newsroom/extensive-gender-discrimination-in-healthcare-access-for-women-in-india/).

Anon. n.d. “Food Security and COVID-19.” Retrieved June 4, 2020j (https://www.worldbank.org/en/topic/agriculture/brief/food-security-and-covid-19).

Anon. n.d. “Health Alert: Panama, LIncreased Movement Restrictions, COVID-19 Situation Report, and Reminders.” Retrieved May 26, 2020k (https://www.osac.gov/Content/Report/c07e97c1-302e-4e36-aed4-18863c5a6fb4).

Anon. n.d. “Hoja de Ruta de La Apertura de Los Comercios, En Yo Me Informo PMA - Portal SERTV.” Retrieved June 4, 2020l (https://sertv.gob.pa/hoja-de-ruta-de-la-apertura-de-los-comercios-en-yo-me-informo-pma/).

Anon. n.d. “Las Lecciones Que Dejó El (Fallido) Intento Del ‘Pico y Género’ En Perú - Cerosetenta.” Retrieved June 1, 2020m (https://cerosetenta.uniandes.edu.co/las-lecciones-que-dejo-el-fallido-intento-del-pico-y-genero-en-peru/).

Anon. n.d. “OxCGRT.” Retrieved June 4, 2020n (https://covidtracker.bsg.ox.ac.uk/stringency-map).

Anon. n.d. “Panama: Set Transgender-Sensitive Quarantine Guidelines | Human Rights Watch.” Retrieved May 26, 2020o (https://www.hrw.org/news/2020/04/23/panama-set-transgender-sensitive-quarantine-guidelines).

Anon. n.d. “Power, Production and Social Reproduction: Human In/Security in the Global … - Google Books.” Retrieved June 5, 2020p (https://books.google.com/books?hl=en&lr=&id=zip-DAAAQBAJ&oi=fnd&pg=PP1&dq=Bakker+Gill+2003+household&ots=Nx6e3GLWTD&sig=DAfU41SLHHh9awmoyFaHmZQ1vF4#v=onepage&q=BakkerGill2003household&f=false).

Anon. n.d. “Socio-Economic Impact of the Ebola Virus Disease in West Africa | UNDP in Africa.” Retrieved June 5, 2020q (https://www.africa.undp.org/content/rba/en/home/library/reports/socio-economic-impact-of-the-ebola-virus-disease-in-west-africa.html).

Anon. n.d. “UK Food Banks Face Record Demand in Coronavirus Crisis | Society | The Guardian.” Retrieved June 4, 2020r (https://www.theguardian.com/society/2020/may/01/uk-food-banks-face-record-demand-in-coronavirus-crisis).

Anon. n.d. “What COVID-19 Tells Us About Gender Inequality in Latin America.” Retrieved June 4, 2020s (https://americasquarterly.org/article/what-covid-19-tells-us-about-gender-inequality-in-latin-america/).

Anon. n.d. “Women Bear Brunt of Coronavirus Economic Shutdown in UK and US | University of Cambridge.” Retrieved June 5, 2020t (https://www.cam.ac.uk/research/news/women-bear-brunt-of-coronavirus-economic-shutdown-in-uk-and-us).

Bandiera, Oriana, Niklas Buehren, Markus P. Goldstein, Imran Rasul, and Andrea Smurra. 2019. “The Economic Lives of Young Women in the Time of EbolaL: Lessons from an Empowerment Program.” 1–80.

Brown, Sally. 2015. “‘They Think It’s All up to the Girls’: Gender, Risk and Responsibility for Contraception.” Culture, Health and Sexuality 17(3):312–25.

Chen, Nanshan, Min Zhou, Xuan Dong, Jieming Qu, Fengyun Gong, Yang Han, Yang Qiu, Jingli Wang, Ying Liu, Yuan Wei, Jia’an Xia, Ting Yu, Xinxin Zhang, and Li Zhang. 2020. “Epidemiological and Clinical Characteristics of 99 Cases of 2019 Novel Coronavirus Pneumonia in Wuhan, China: A Descriptive Study.” The Lancet 395(10223):507–13.

Davies, Sara E. and Belinda Bennett. 2016. “A Gendered Human Rights Analysis of Ebola and Zika: Locating Gender in Global Health Emergencies.” International Affairs 92:1041–60.

Diniz, Debora and Diane R. Grosklaus Whitty. n.d. Zikal: From the Brazilian Backlands to Global Threat.

Elson, Diane. 1994. “People, Development and International Financial Institutions: An Interpretation of the Bretton Woods System.” Review of African Political Economy 21(62):511–24.

Goldenberg, Shira M., Jill Chettiar, Paul Nguyen, Sabina Dobrer, Julio Montaner, and Kate Shannon. 2014. “Complexities of Short-Term Mobility for Sex Work and Migration among Sex Workers: Violence and Sexual Risks, Barriers to Care, and Enhanced Social and Economic Opportunities.” Journal of Urban Health 91(4):736–51.

Hanson, Susan. 2010. “Gender and Mobility: New Approaches for Informing Sustainability.” Gender, Place and Culture 17(1):5–23.

Hapke, Holly M. and Devan Ayyankeril. 2004. “Gender, the Work-Life Course, and Livelihood Strategies in a South Indian Fish Market.” Gender, Place and Culture 11(2):229–56.

Harman, Sophie. 2016. “Ebola, Gender and Conspicuously Invisible Women in Global Health Governance.” Third World Quarterly 37(3):524–41.

Jejeebhoy, Shireen J. and Zeba A. Sathar. 2004. “Women’s Autonomy in India and Pakistan: The Influence of Religion and Region.” Population and Development Review 27(4):687–712.

Jejeebhoy, Shireen J. and Zeba A. Sathar. n.d. Women’s Autonomy in India and Pakistan: The Influence of Religion and Region.

Lanier, Christina and Michael O. Maume. 2009. “Intimate Partner Violence and Social Isolation across the Rural/Urban Divide.” Violence Against Women 15(11):1311–30.

Lee, Ellie and Elizabeth Frayn. 2008. “The ‘Feminisation’ of Health.” Pp. 115–33 in A Sociology of Health. SAGE Publications Inc.

Liu, Shiwei, Mei Zhang, Ling Yang, Yichong Li, Limin Wang, Zhengjing Huang, Linhong Wang, Zhengming Chen, and Maigeng Zhou. 2017. “Prevalence and Patterns of Tobacco Smoking among Chinese Adult Men and Women: Findings of the 2010 National Smoking Survey.” Journal of Epidemiology and Community Health 71(2):154–61.

Mahase, Elisabeth. 2020. “Covid-19: EU States Report 60% Rise in Emergency Calls about Domestic Violence.” BMJ (Clinical Research Ed.) 369:m1872.

Mandel, Jennifer L. 2004. “Mobility Matters: Women’s Livelihood Strategies in Porto Novo, Benin.” Gender, Place and Culture 11(2):257–87.

Marmot, Michael. 2005. “Social Determinants of Health Inequalities.” Lancet 365(9464):1099–1104.

Mumtaz, Zubia and Sarah Salway. 2005. “‘I Never Go Anywhere’: Extricating the Links between Women’s Mobility and Uptake of Reproductive Health Services in Pakistan.” Social Science and Medicine 60(8):1751– 65.

Osamor, Pauline E. and Christine Grady. 2016. “Women’s Autonomy in Health Care Decision-Making in Developing Countries: A Synthesis of the Literature.” International Journal of Women’s Health 8:191–202.

Phimister, Angus, Sonya Krutikova, Lucy Kraftman, Christine Farquharson, Monica Costa Dias, Sarah Cattan, Alison Andrew, and Almudena Sevilla. 2020. How Are Mothers and Fathers Balancing Work and Family under Lockdown?

Pronyk, Paul M., James R. Hargreaves, Julia C. Kim, Linda A. Morison, Godfrey Phetla, Charlotte Watts, Joanna Busza, and John DH Porter. 2006. “Effect of a Structural Intervention for the Prevention of Intimate-Partner Violence and HIV in Rural South Africa: A Cluster Randomised Trial.” Lancet 368(9551):1973–83.

Roberts, Adrienne. 2013. “Financing Social Reproduction: The Gendered Relations of Debt and Mortgage Finance in Twenty-First-Century America.” New Political Economy 18(1):21–42.

Sager, Tore. 2016. “Gendered Mobility: A Case Study of Non-Western Immigrant Women in Norway.” 89–118.

Samari, Goleen and Anne R. Pebley. 2018. “Longitudinal Determinants of Married Women’s Autonomy in Egypt.” Gender, Place and Culture 25(6):799–820.

Sjoberg, Laura. 2016. “What, and Where, Is Feminist Security Studies?” Journal of Regional Security 11(2):143–61.

Smith, D. E. 1990. The Conceptual Practices of Power: A Feminist Sociology. Vol. 24. University of Toronto Press.

Smith, Julia. 2019. “Overcoming the ‘Tyranny of the Urgent’: Integrating Gender into Disease Outbreak Preparedness and Response.” Gender and Development 27(2):355–69.

Thornton, Jacqui. 2020. “Covid-19: A&E Visits in England Fall by 25% in Week after Lockdown.” BMJ (Clinical Research Ed.) 369:m1401.

Wenham, Clare, João Nunes, Gustavo Correa Matta, Carolina de Oliveira Nogueira, Polyana Aparecida Valente, and Denise Nacif Pimenta. 2020. “Gender Mainstreaming as a Pathway for Sustainable Arbovirus Control in Latin America” edited by C. I. Brodskyn. PLOS Neglected Tropical Diseases 14(2):e0007954.

Wenham, Clare, Julia Smith, and Rosemary Morgan. 2020. “COVID-19: The Gendered Impacts of the Outbreak.” The Lancet 395(10227):846–48.

